# Exposure to cough aerosols and development of pulmonary COVID-19

**DOI:** 10.1101/2020.06.03.20121004

**Authors:** Koen Vanden Driessche, Jeremy Nestele, Jeroen Grouwels, Els LIM Duval

## Abstract

Half a year after the emergence of COVID-19, research is still going on to gain insight in the importance of different SARS-CoV-2 transmission routes and their impact on the clinical picture of COVID-19. Our findings suggest that coughing is not as important for transmission as initially anticipated and we discuss the potentially important role for loud conversation as a driver for transmission.

**Disclosure:** None of the authors have any financial associations or other possible conflicts of interest to declare.

## Introduction

Six months after the emergence of SARS-CoV-2 the relative importance of its different transmission routes remains to be clarified. Respiratory viruses like SARS-CoV-2 are normally transmitted by contact (direct and/or indirect contact) or when the virus travels on tiny droplets released into the air. That’s why, in the beginning of the COVID-19 pandemic it was anticipated that SARS-CoV-2 transmission started with a sneeze or cough, similarly to SARS-CoV-1. Cough aerosols contain a mix of droplets and airborne particles. Droplets remain larger than 5-10 micron while quickly falling to the ground. Airborne particles, formally called “droplet nuclei”, originate from droplets that become so small while evaporating that they remain suspended in the air. The *World Health Organization* believes that SARS-CoV-2 normally does not transmit through droplet nuclei, only through droplets. Weather on droplets or droplet nuclei or a combination of both, a recent hospital study is suggestive for important SARS-CoV-2 transport through the air, demonstrated by high ratios of positive air and floor samples (1).

We previously reported that surgical masks worn by cystic fibrosis patients while coughing, not only stop droplets, but also block the generation of droplet nuclei containing viable *Pseudomonas* (2). While a lot of air, if not most, escapes through the gaps between face and mask when coughing, the infectious particles did not (2–3). Our findings were confirmed by another group and very recently a study with seasonal coronaviruses reported similar pilot results (4–5). The behavior of virus-laden droplets and droplet nuclei in aerosols, depends on the aerodynamic size of these carrier particles, not on the size of the virus itself.

We hypothesized that the SARS-CoV-2 route of transmission is a disease severity determinant and that most (more than 70%) patients with severe pulmonary COVID-19 were exposed to cough aerosols. Based on this, one could expect a significant effect of mass masking on COVID-19 morbidity and mortality.

## Methods

After explaining the COVID-19 incubation period, we asked one question to patients that were almost 100% certain which person infected them: “Was coughing a symptom of this person?”. Confidence intervals (CI) for proportions were calculated using MedCalc^®^ (MedCalc Software, Ostend, Belgium). The study received a review exemption from the Ethics Committee of Antwerp University Hospital in Belgium.

## Results

We received thirty-eight filled “ONE-questionnaires” between March 18 and April 17. Only 9 out of 25 hospitalized patients requiring supplemental oxygen (36%; 95% CI 18-57%) got infected by somebody who coughed (Figure 1). This result does not support the hypothesis that exposure to cough aerosols is required (meaning observed in over 70% of the patients) to develop severe pulmonary COVID-19. Overall, only 14 out of 38 COVID-19 patients (37%; 95% CI 22-54%) were infected by someone who had coughing as a symptom.

**Figure 1.**
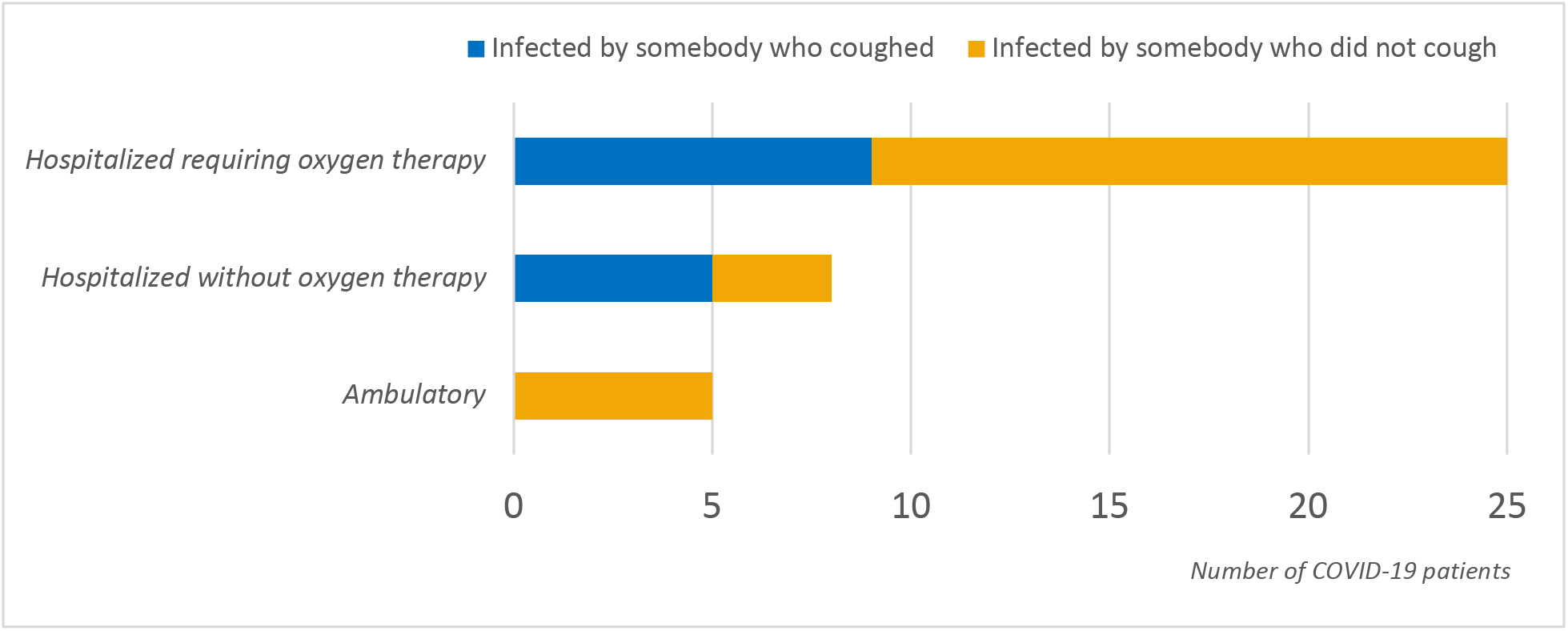
Results of the ONE-questionnaire

## Discussion

Participating patients were only asked to answer one question, which was both a strength and a limitation of our study. As with any case-control study, there is a risk of recall bias and misclassification: patients may not remember coughing as a symptom of the person who infected them. Nevertheless, a lot of the transmission did not seem to involve coughing at all. Alternatively, other respiratory maneuvers like sneezing, talking or breathing could have generated SARS-CoV-2 aerosols. When talking, vibration of the vocal cords generates 2.5-micron droplet nuclei (6). Ten seconds saying “aah” produces a comparable number of droplet nuclei as coughing for 10 seconds (7). The vocal cords are being lubricated by saliva, which contains large amounts of SARS-CoV-2 in most COVID-19 patients (8). During singing or talking with a loud voice, up to 50 times more droplet nuclei are being generated compared to talking with a quiet voice (9–10). Researchers also noticed that 1 in 5 people released 10 times more particles during talking than their peers for reasons not yet understood. Bringing all this information together, one could expect COVID-19 superspreader events in bars and clubs with loud music, choirs, and crowded markets. Lately these are increasingly being reported, in both non-scientific and scientific press. One could even argue that talking with a loud voice to hearing impaired elderly, could have contributed to the high COVID-19 incidence in nursing homes. Droplets have enough mass and speed to get impacted in a mask when coughing, even the small ones that would otherwise evaporate to droplet nuclei (2). Large droplets also impact in a mask during speech, but there is no guarantee that masks prevent small droplets from evaporating to droplet nuclei when just talking or breathing. According to Newton’s second law of motion (F=m.a), less force is required to deflect the slowly moving particles during speech or breathing, which could make it easier for them to follow airflows around the edge of a mask.

After droplet or contact infection of the upper respiratory tract, SARS-CoV-2 could spread to the lower airways through a process of auto-inoculation, which is an alternative hypothesis for the pathogenesis of lung lesions in COVID-19 that would not involve direct inhalation of virus-laden particles. Another possible mechanism could involve hematogenous spread to the lungs.

In conclusion, our study could not confirm that exposure to cough aerosols is required for developing severe pulmonary COVID-19. Further research is needed to look into transmission routes of SARS-CoV-2 and the preventive role of face masks or even “voice etiquette”.

## Data Availability

All original data are filed at Antwerp University Hospital in Belgium. Please contact us for access.

## Authors’ contributions

KVD and ED conceived the study, analyzed the data, and drafted the manuscript. JG and JN collected the data and reviewed the manuscript. All authors approved the final version of the article.

